# Proteomic Pathways across Ejection Fraction Spectrum in Heart Failure: an EXSCEL Substudy

**DOI:** 10.1101/2023.05.16.23288273

**Authors:** Anthony E. Peters, Maggie Nguyen, Jennifer B. Green, Ewan R Pearson, John Buse, Harald Sourij, Adrian F. Hernandez, Naveed Sattar, Rury R. Holman, Robert J. Mentz, Svati H. Shah

**Author notes:** **Address for Correspondence**: Svati H. Shah, Duke Molecular Physiology Institute, 300 N. Duke Street, Durham NC 27710.

## Abstract

**Background:** Ejection fraction (EF) is a key component of heart failure (HF) classification, including the increasingly codified HF with mildly reduced EF (HFmrEF) category. However, the biologic basis of HFmrEF as an entity distinct from HF with preserved EF (HFpEF) and reduced EF (HFrEF) has not been well characterized.

**Methods:** The EXSCEL trial randomized participants with type 2 diabetes (T2DM) to once-weekly exenatide (EQW) vs. placebo. For this study, profiling of ∼5000 proteins using the SomaLogic SomaScan platform was performed in baseline and 12-month serum samples from N=1199 participants with prevalent HF at baseline. Principal component analysis (PCA) and ANOVA (FDR p<0.1) were used to determine differences in proteins between three EF groups, as previously curated in EXSCEL (EF>55% [HFpEF], EF 40-55% [HFmrEF], EF<40% [HFrEF]). Cox proportional hazards was used to assess association between baseline levels of significant proteins, and changes in protein level between baseline and 12-month, with time-to-HF hospitalization. Mixed models were used to assess whether significant proteins changed differentially with exenatide vs. placebo therapy.

**Results:** Of N=1199 EXSCEL participants with prevalent HF, 284 (24%), 704 (59%) and 211 (18%) had HFpEF, HFmrEF and HFrEF, respectively. Eight PCA protein factors and 221 individual proteins within these factors differed significantly across the three EF groups. Levels of the majority of proteins (83%) demonstrated concordance between HFmrEF and HFpEF, but higher levels in HFrEF, predominated by the domain of extracellular matrix regulation, *e.g.* COL28A1 and tenascin C [TNC]; p<0.0001. Concordance between HFmrEF and HFrEF was observed in a minority of proteins (1%) including MMP-9 (p<0.0001). Biologic pathways of epithelial mesenchymal transition, ECM receptor interaction, complement and coagulation cascades, and cytokine receptor interaction demonstrated enrichment among proteins with the dominant pattern, *i.e.* HFmrEF-HFpEF concordance. Baseline levels of 208 (94%) of the 221 proteins were associated with time-to-incident HF hospitalization including domains of extracellular matrix (COL28A1, TNC), angiogenesis (ANG2, VEGFa, VEGFd), myocyte stretch (NT-proBNP), and renal function (cystatin-C). Change in levels of 10 of the 221 proteins from baseline to 12 months (including increase in TNC) predicted incident HF hospitalization (p<0.05). Levels of 30 of the 221 significant proteins (including TNC, NT-proBNP, ANG2) were reduced differentially by EQW compared with placebo (interaction p<0.0001).

**Conclusions:** In this HF substudy of a large clinical trial of people with T2DM, we found that serum levels of most proteins across multiple biologic domains were similar between HFmrEF and HFpEF. HFmrEF may be more biologically similar to HFpEF than HFrEF, and specific related biomarkers may offer unique data on prognosis and pharmacotherapy modification with variability by EF.

## INTRODUCTION

Left ventricular ejection fraction (LVEF) phenotype, now including HF with mildly reduced EF (HFmrEF), is a key component of classifying patients with heart failure (HF) and guiding implementation of therapies.^1, 2^ Yet, the biologic basis of HFmrEF as an entity distinct from HF with preserved EF (HFpEF) and HF with reduced EF (HFrEF) has not been well characterized. There is accumulating evidence for different biological signatures underlying HFpEF as compared with HFrEF,^3–5^ but HFmrEF remains a less well-defined entity. Data on HFmrEF as comprising a distinct pathophysiology or representing simply a transition between HFrEF and HFpEF are mixed.^6^ Prior data has suggested that HFmrEF has some similarities to HFrEF with regards to HF etiology (e.g. more commonly ischemic), outcomes, and response to therapy,^7–11^ although these data are inconsistent with some recent studies indicating more clinical similarities between HFpEF and HFmrEF.^12^

Molecular profiling can help evaluate underlying biological basis of disease while simultaneously identifying clinically relevant biomarkers. Notably, proteomics studies including HFmrEF have shown unique, perhaps intermediary, signatures in HFmrEF, but have been limited by small sample sizes, focus on a smaller set of proteins, and/or lack of longitudinal biomarker data.^4, 5, 13, 14^ At a clinical level, there is an increasing movement to advance beyond LVEF in phenotyping patients with HF due the limited relationship between LVEF and pathophysiology, outcomes, and treatment response.^15–17^ An improved understanding of the biological correlates of LVEF phenotypes and related biomarkers in HF could inform patient classification and management. Thus, to address these interconnected gaps in the literature, we used proteomics applied to a large clinical trial database to define molecular processes across the EF spectrum in HF and investigated their role as prognostic biomarkers of incident hospitalization for HF (hHF) outcomes.

## METHODS

### Study Population

The Exenatide Study of Cardiovascular Event Lowering (EXSCEL) evaluated the effects of once-weekly exenatide (EQW) in adults with type 2 diabetes mellitus (T2DM). Design, baseline characteristics, and primary results have been published.^18–20^ The EXSCEL study population was enriched for participants with history of prior CV events including previous coronary, cerebrovascular, or peripheral vascular events or stenosis and also included a primary prevention group.^19^ Biospecimens were collected at enrollment in a subset of EXSCEL participants (N=5668, 38.4% of the overall trial population).

For this study, participants with evidence of prevalent baseline HF who had consented for and had biospecimens collected were included. Presence or absence of history of clinical HF was captured upon enrollment into the trial. Clinical HF status at baseline was prospectively recorded by the clinician-investigator based on all available clinical data including patients’ signs/symptoms and objective measures such as echocardiography and biomarker data (eg, natriuretic peptide levels).^21^ Participants with prevalent HF were further stratified across three EF groups as defined and curated in the parent EXSCEL trial as follows: preserved ejection fraction (HFpEF, EF>55%), mildly reduced ejection fraction (HFmrEF, EF 40-55%), and reduced ejection fraction (HFrEF, EF<40%). Incident HF events were defined as hHF as the primary reason for hospitalization. A blinded, independent clinical events classification committee adjudicated all the components of the primary composite outcome and secondary outcomes including hHF.

### Proteomic profiling

Profiling of ∼5000 proteins was performed in frozen serum collected at baseline (enrollment) and 12 month follow-up. Profiling was performed utilizing the SomaScan assay (SomaLogic Inc., Boulder, USA). The design and performance characteristics of this assay have been previously described.^22, 23^ This assay uses DNA-based binding reagents (modified aptamers) to quantify, with high specificity, the availability of binding epitopes on plasma proteins^22, 24^, enabling high throughput relative quantification of thousands of proteins in small amounts of sample. Samples were run in three dilutions to ensure dynamic range in serum. Eleven control replicates from three control lots were included in each 96-sample plate and five calibrator replicates per run were used with a reference standard. Three QC replicates were also included per run with a reference standard to evaluate the accuracy of the assay after data standardization. Prior studies have established the specificity of the majority of the SomaScan reagents using affinity-capture experiments and orthogonal means (i.e., presence of *cis* genetic variants and validation by mass spectrometry).^25, 26^

### Statistical analysis

Proteomics data first underwent quality control (QC) procedures and standard normalization^27^ using adaptive median normalization by maximum likelihood, scaling the total fluorescence from the experimental sample based on point and variance estimates from a population control set as previously done. This procedure identified 5.1% of samples for having normalization scale factors outside recommended ranges and were removed from analysis. Unsupervised principal component analysis (PCA) with varimax rotation was performed on 4979 proteins (baseline and 12-month samples) passing QC measures as a means of dimensionality reduction to reduce the burden of multiple comparisons and identify potential shared biologic pathways. The Kaiser criterion (eigenvalue >1) was used to identify PCA protein factors for inclusion in analysis. Each PCA factor is a weighted sum of all proteins for each participant. Proteins with an absolute value factor load ≥0.4 were considered as heavily loaded and thus composing a given factor and were analyzed as individual proteins for significant PCA factors. PCA factors with only one protein heavily loaded in a given factor were not analyzed as a factor but instead as the individual heavily loaded protein.

For discovery analyses, ANOVA was used to determine differences in PCA protein factor levels across the three EF groups (HFpEF, HFmrEF and HFrEF) and adjusted for multiple comparisons using the Benjamini Hochberg false discovery rate [FDR] p<0.1 at level of factors (**Supplemental Figure 1**). Individual proteins heavily loaded on PCA factors significant from the ANOVA were then analyzed in subsequent sensitivity analyses (nominal significance i.e. p<0.05) as follows: (1) ANOVA across EF groups; (2) of proteins significant in ANOVA, unadjusted pairwise comparisons using t-tests with pooled SD between the three EF groups; (3) significant individual proteins were then assessed in pairwise multivariable model adjusted for age, sex, race, hypertension, hyperlipidemia, obesity by enrollment body mass index (BMI ≥ 30 kg/m^2^), and enrollment hemoglobin A1C; (4) Cox proportional hazard models were constructed to assess for relationship between baseline protein and change in protein level (baseline to 12 month) with time-to-incident hHF; for proteins violating the proportional hazards assumption, a parametric accelerated failure time (AFT) model was constructed for comparison; and (5) the effect of EQW on protein levels was assessed using linear mixed effects models inclusive of terms for treatment arm (EQW vs. placebo), protein timepoint (baseline, 12 months) and an interaction term (treatment*timepoint). For all incident HF hospitalization analyses, only events occurring after 12-month follow-up were included.

Based on results from pairwise multivariable models (sensitivity analysis #3), individual proteins were classified into patterns for being concordant or discordant between EF groups (i.e. based on nominal p-value, proteins not significantly different between two EF groups were considered concordant between those groups; proteins significantly different between two EF groups were considered discordant) and by whether protein levels were significantly higher/lower in one EF group (or increasing/decreasing significantly across groups).

Overrepresentation analysis (ORA) using the Molecular Signatures Database (MSigDB) was performed to highlight biologic pathways enriched among individual proteins, starting with those with multivariable ANOVA p-value <0.05 among EF groups, and narrowing to those with 1) concordant levels in HFmrEF and HFpEF (multivariable pairwise p-value ≥ 0.05) and, separately, 2) concordant levels in HFmrEF and HFrEF (multivariable pairwise p-value ≥ 0.05). ORA was conducted using a hypergeometric test of these specific proteins on a background of all tested proteins. A pathway was considered significantly enriched if its nominal p-value ≤ 0.05. Two gene collections (Hallmark [50 gene sets] and Canonical Pathways derived from the KEGG pathway database [CP:KEGG; 186 gene sets]) were utilized. Primary biological domains (i.e. extracellular matrix/space regulation, endothelial-mesenchymal transition, angiogenesis, etc.) for this analysis were defined by presence in a combination of multiple sources including ‘Biological Processes’ and Cellular Components’ of Gene Ontology, biological pathways defined by the Molecular Signatures Database (MSigDB), and several HF-biomarker literature sources.^5, 28–31^

## RESULTS

### Baseline characteristics and differences between EF groups

Among EXSCEL participants with biospecimens available (N=5668, 38.4% of total EXSCEL population), N=1199 (21.2%) of individuals had prevalent HF. Of these, N=284 (24%) had HFpEF, N=704 (59%) had HFmrEF, and N=211 (18%) had HFrEF. Baseline characteristics of the study population overall and by EF group are included in **Table 1**. Participants with HFmrEF had an overall intermediate demographic and clinical profile compared to participants with HFrEF or HFpEF as follows: participants with HFmrEF were 71% male (compared to 78% of HFrEF and 64% of HFpEF), 90% white (compared to 83% of HFrEF and 95% of HFpEF), and 66% enrolled in Europe (compared to 47% for HFrEF and 82% for HFpEF). Participants with HFmrEF were 41% non-smokers (compared to 60% of HFpEF and 38% of HFrEF) and had an intermediate burden of prior cardiovascular events and coronary disease at baseline, compared to participants with HFrEF or HFpEF (**Table 1**). Participants with HFrEF demonstrated higher rates of HF hospitalization, cardiovascular mortality, and all-cause mortality, compared to those with HFmrEF or HFpEF (**Supplemental Table 1**).

**Table 1.** Baseline characteristics of participants by ejection fraction group.

|  | HFpEF (n=284) | HFmrEF (n=704) | HFrEF (n=211) | p-value |
| --- | --- | --- | --- | --- |
| Age | 63.96 (8.91) | 62.63 (9.12) | 63.95 (9.45) | 0.047 |
| Male sex | 181 (63.7) | 500 (71.0) | 164 (77.7) | 0.003 |
| Race |  |  |  | 0.001 |
| White | 274 (96.5%) | 630 (89.5) | 175 (82.9%) |  |
| Black | 4 (1.4%) | 12 (1.7%) | 7 (3.3%) |  |
| Asian | 5 (1.8%) | 26 (3.7) | 12 (5.7%) |  |
| Hispanic | 1 (0.4%) | 31 (4.4%) | 16 (7.6%) |  |
| Indian (American) or Alaska Native | 0 (0.0%) | 3 (0.4%) | 1 (0.5%) |  |
| Native Hawaiian or Other Pacific Islander | 0 (0.0%) | 2 (0.3%) | 0 (0.0%) |  |
| Region |  |  |  | <0.001 |
| Asia Pacific | 7 (2.5%) | 28 (4.0%) | 12 (5.7%) |  |
| Europe | 234 (82.4%) | 466 (66.2%) | 99 (46.9%) |  |
| Latin America | 0 (0.0%) | 20 (2.8%) | 17 (8.1%) |  |
| North America | 43 (15.1%) | 190 (27.0%) | 83 (39.3%) |  |
| Smoking status |  |  |  | <0.001 |
| Current | 26 (9.2%) | 86 (12.2%) | 21 (10.0%) |  |
| Former | 88 (31.0%) | 327 (46.4%) | 109 (51.7%) |  |
| Never | 170 (59.9%) | 291 (41.3%) | 81 (38.4%) |  |
| Medical history at baseline |  |  |  |  |
| Prior Cardiovascular Event at Randomization | 234 (82.4%) | 635 (90.2%) | 196 (92.9%) | <0.001 |
| Hx of MI | 122 (43.0%) | 414 (58.8%) | 157 (74.4%) | <0.001 |
| Prior PCI | 52 (24.3%) | 242 (46.9%) | 62 (42.5%) | <0.001 |
| Prior CABG | 42 (19.6%) | 134 (26.0%) | 54 (37.0%) | 0.001 |
| Hx of Cerebrovascular Disease | 61 (21.5%) | 118 (16.8%) | 25 (11.8%) | 0.018 |
| Hx of Peripheral Arterial Disease | 58 (20.4%) | 105 (14.9%) | 37 (17.5%) | 0.10 |
| Hypertension | 266 (93.7%) | 661 (93.9%) | 188 (89.1%) | 0.05 |
| Hyperlipidemia | 208 (73.2%) | 614 (87.2%) | 175 (82.9%) | <0.001 |
| Duration of Diabetes at BL | 13.00 (9.34) | 12.67 (8.22) | 12.45 (8.52) | 0.76 |
| Depression | 10 (4.7%) | 44 (8.5%) | 23 (15.8%) | 0.001 |
| Medications at baseline |  |  |  |  |
| Insulin | 142 (50.0%) | 334 (47.4%) | 115 (54.5%) | 0.19 |
| Metformin | 191 (67.3%) | 504 (71.6%) | 124 (58.8%) | 0.002 |
| Sulfonylurea | 100 (35.2%) | 256 (36.4%) | 54 (25.6%) | 0.014 |
| Thiazolidinedione | 9 (3.2%) | 18 (2.6%) | 2 (0.9%) | 0.26 |
| Non-sulfonylurea | 2 (0.7%) | 5 (0.7%) | 0 (0.0%) | 0.47 |
| Alpha-glucosidase | 0 (0.0%) | 9 (1.3%) | 2 (0.9%) | 0.16 |
| GLP-1 Analogues | 284 (100.0%) | 704 (100.0) | 211 (100.0%) | NA |
| SGLT-2 Inhibitors | 0 (0.0%) | 1 (0.1%) | 0 (0.0%) | 0.71 |
| Aspirin | 221 (77.8%) | 526 (74.7%) | 160 (75.8%) | 0.59 |
| Vitamin K antagonist | 21 (7.4%) | 90 (12.8%) | 43 (20.4%) | <0.001 |
| Statin | 228 (80.3%) | 596 (84.7%) | 179 (84.8%) | 0.21 |
| Ezetimibe | 10 (3.5%) | 44 (6.2%) | 17 (8.1%) | 0.091 |
| Fibrate | 12 (4.2%) | 60 (8.5%) | 11 (5.2%) | 0.031 |
| Niacin | 9 (3.2%) | 14 (2.0%) | 4 (1.9%) | 0.49 |
| Fish oil | 13 (4.6%) | 56 (8.0%) | 21 (10.0%) | 0.063 |
| Chronic Non-Steroidal Anti-Inflammatory Drugs | 8 (2.8%) | 27 (3.8%) | 12 (5.7%) | 0.26 |
| Hormone Replacement Therapy | 6 (2.1%) | 18 (2.6%) | 13 (6.2%) | 0.016 |
| Body Mass Index | 33.85 (6.71) | 34.10 (6.50) | 32.85 (6.45) | 0.055 |
| Systolic BP | 136.62 (15.40) | 135.16 (15.01) | 128.43 (17.01) | <0.001 |
| Diastolic BP (mean (SD)) | 79.75 (9.68) | 77.75 (10.31) | 75.77 (10.79) | <0.001 |
| HbA1c (%) (mean (SD)) | 8.25 (0.98) | 8.13 (0.96) | 8.05 (0.97) | 0.053 |
| Creatinine (mg/dl) (mean (SD)) | 1.04 (0.29) | 1.02 (0.28) | 1.15 (0.34) | <0.001 |
| eGFR (mean (SD)) | 76.76 (20.47) | 79.20 (20.69) | 72.07 (21.94) | <0.001 |

Values are mean (SD) or n (%)

### Protein dimensionality reduction and association between protein factors with EF groups

PCA with varimax rotation of N=4979 proteins resulted in 852 protein factors; 120 factors had multiple heavily loaded proteins and 732 factors had just a single protein heavily loaded on the factor (**Supplemental Table 2**). Factors heavily loaded with a single protein were analyzed instead as the individual protein. In discovery analyses across all protein factors/proteins, ANOVA (FDR adjusted for multiple comparisons considering 852 comparisons) across the three EF groups identified three PCA protein factors (factors 5, 95, and 625) and five individual proteins that were significantly different (FDR<0.1) across EF groups (**Supplemental Table 3)**. The three PCA protein factors were comprised of 249 (factor 5), six (factor 95) and five (factor 625) heavily loaded proteins respectively. Proteins most heavily loaded in the three PCA factors included ganglioside GM2 activator, neuroblastoma suppressor of tumorigenicity 1, and cystatin-C (factor 5); tenascin-C and chymotrypsin-like elastase family member 1 (factor 95); and matrix metalloproteinase-9, cysteine-rich secretory protein LCCL domain-containing 2, and prokineticin-2 (factor 625). The five individual proteins were endothelin-2, cytokine receptor-like factor 1, keratin, type 1 cytoskeletal 16, properdin, and troponin I. Of note, NT-proBNP was loaded on factor 5 (factor loading 0.487) with an individual protein nominal p-value <0.001.

In analyses of individual proteins loaded on these PCA protein factors, 247 individual proteins remained significantly associated with EF groups. The majority of these proteins (n=221, 90%) remained significant in multivariable models (197 from PCA factors and five from PCA factors with only one protein heavily loaded) with strongest results for NTpro-BNP, cystatin-C, and collagen alpha-1(XXVIII) chain (COL28A1).

To further understand the biologic pathways represented by these proteins, overrepresentation analyses (ORA) were performed which revealed several pathways (**Table 2**) to be significantly enriched among proteins that were concordant between HFmrEF and HFpEF (587 of 4979 proteins, translating to 568 of 4755 genes, all with nominal p<0.05) including pathways of epithelial-mesenchymal transition (nominal p<0.001, 34 genes including those associated with IGF binding proteins, TNC, TNF receptors, and VEGFa), ECM receptor interaction, complement and coagulation cascades, cytokine receptor interaction, cell adhesion molecules, and angiogenesis. ORA demonstrated fewer pathways (**Supplemental Table 4**) to be significantly enriched among proteins with similar expression levels between HFmrEF and HFrEF (116 proteins among 4755, all with nominal p<0.05). These pathways included systemic lupus erythematosus, starch and sucrose metabolism, and folate biosynthesis.

**Table 2.** Overrepresentation analysis for proteins with HFmrEF-HFpEF concordance.

|  | Pathway size | Significant genes in pathway | Nominal p-value | Select genes |
| --- | --- | --- | --- | --- |
| Epithelial-mesenchymal transition (EMT) | 143 | 34 | <0.001 | IGFBPs, TNC, VEGFa |
| Cell adhesion molecules (CAMs) | 69 | 17 | 0.002 | CADMs, CD8a |
| Complement and coagulation cascades | 61 | 14 | 0.01 | CD46, CD55 |
| Extracellular matrix receptor interaction | 47 | 11 | 0.02 | COL6A2, TNC |
| Apical surface | 23 | 7 | 0.02 | CD160, IL2RB |
| Cytokine and cytokine receptor interaction | 228 | 37 | 0.03 | EGFR, TNFRs, VEGFa |
| Angiogenesis | 28 | 7 | 0.04 | FGFR1, VEGFa |
| Estrogen response early | 66 | 13 | <0.05 | IGFBPs, TFF3 |
| Axon guidance | 93 | 17 | <0.05 | EFNA2, NTN4 |
| IL6, JAK, STAT3, signaling | 73 | 14 | <0.05 | IL15RA, TNFRs |

### Protein patterns across EF groups

Patterns across EF groups were defined based on nominal significance (p<0.05) of multivariable pairwise comparisons between groups. Overall, 193 (87%) of these proteins were differentially elevated in one EF group, while six (2.7%) of these proteins demonstrated a graded increasing/decreasing pattern across EF groups.

The dominant pattern across groups was of higher levels of proteins in HFrEF as compared to both HFmrEF and HFpEF, with concordance in protein levels in HFmrEF as compared with HFpEF (n=184 proteins, 82.9%; **Central Illustration**, **Figure 4, Supplemental Figure 2**). This pattern included previously established biomarkers of troponin T, troponin I, and cystatin-C, as well as proteins residing in multiple biologic domains including extracellular matrix/space regulation (e.g. COL28A1, TNC), angiogenesis (e.g. ANG2, VEGFa, VEGFd), humoral immune response (e.g. B2M), inflammation (e.g. TNF ligands/receptors), and growth factor response/regulation (e.g. EGFR, IGFBP2). All individual protein multivariable ANOVA across EF groups nominal p<0.001. Three proteins (1.35%) (MMP-9, cysteine-rich secretory protein LCCL domain-containing 2, and prokineticin-2) demonstrated a pattern with significantly different levels in HFpEF as compared with both HFrEF and HFmrEF (**Supplemental Figure 4**). Three proteins (1.35%) (selenoprotein W, CD48 antigen, programmed cell death 1 ligand 2) demonstrated a pattern with different levels in HFmrEF as compared with both HFrEF and HFpEF, with each protein with lower levels in HFmrEF compared to other groups (**Supplemental Figure 3**). Six proteins (2.4%) showed a steadily increasing pattern of protein levels across EF groups including one protein (properdin) with higher levels in HFpEF, intermediate in HFmrEF and lowest in HFrEF and 5 proteins with higher levels in HFrEF, intermediate in HFmrEF and lowest in HFpEF: NTpro-BNP, hepatitis A virus cellular receptor 2 (TIMD3), neutrophil gelatinase-associated lipocalin (lipocalin 2), kallikrein-11, and transcobalamin-1 (holo-TC 1) (all multivariable nominal ANOVA p<0.0001, **Supplemental Figure 5**).

### Association between identified proteins and incident hHF risk

Baseline levels of the majority of individual proteins significantly associated with EF group (n=213/221, 96%) also demonstrated association with time-to-incident hHF in the multivariable analysis (**Figure 1**), as expected, given higher event rates in participants with HFrEF (**Supplemental Table 1**); 22 of these 213 proteins demonstrated violation of the PH assumption in the multivariable Cox model and were evaluated by parametric AFT model which showed concordant results with the primary Cox model and thus primary Cox model analyses are included here (**Supplemental Table 5**). Proteins whose baseline levels were associated with time to incident hHF included COL28A1 (multivariable HR 13.97 [95% CI 8.89-21.94], nominal p<0.0001), cystatin-C (HR 13.54 [95% CI: 8.25-22.22], p<0.0001), angiopoietin-2 (HR 8.9 [95% CI: 6.6-12.0], p<0.0001), TNC [HR 8.87 [95% CI: 6.55-12.02], p<0.0001), tumor necrosis factor receptor superfamily member 1A (HR 6.54 [95% CI: 4.30-9.93], p<0.0001), and NT-proBNP (HR 3.13 [95% CI: 2.68-3.67], p<0.0001). Notably, each of these proteins had demonstrated a pattern of higher levels in HFrEF and similar levels in HFmrEF/HFpEF, except for NTpro-BNP, which after multivariable adjustment, showed a graded increasing pattern across the three EF groups.

**Figure 1:**
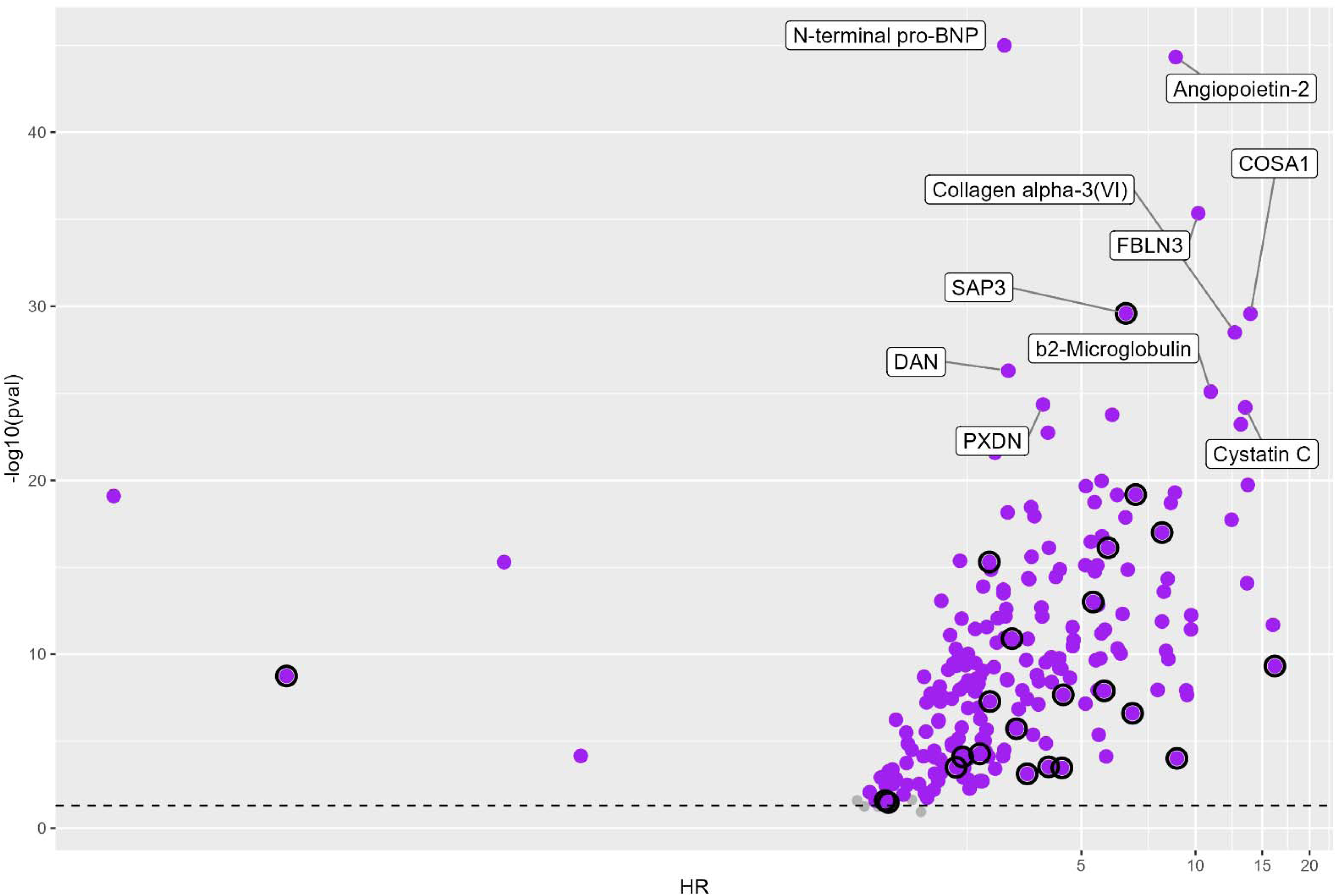
Volcano plot of association between baseline protein expression and risk of heart failure hospitalization in multivariable analysis. Black circles indicate proteins violating PH assumption.

Given the potential clinical utility of understanding how changes in these biologic pathways may presage HF development, we then examined whether change in protein levels from baseline to 12 months was associated with time-to-incident hHF. Ten of these proteins were associated with time-to-incident hHF (i.e. events after 12 month follow-up) including transmembrane emp24 domain-containing protein 10 (multivariable HR 4.55 [2.00-10.55], p=0.0004), TNC (HR 2.02 [1.08-3.77], p=0.03), and epidermal growth factor receptor (HR 0.09 [0.02-0.61], p=0.01) (**Figure 2**). Two proteins (transmembrane emp24 domain-containing protein 10 and TNC) demonstrated higher risk with an increase in mean protein levels, with the other proteins showing higher risk with a decrease in mean protein levels (**Supplemental Figures 7 and 8**).

**Figure 2:**
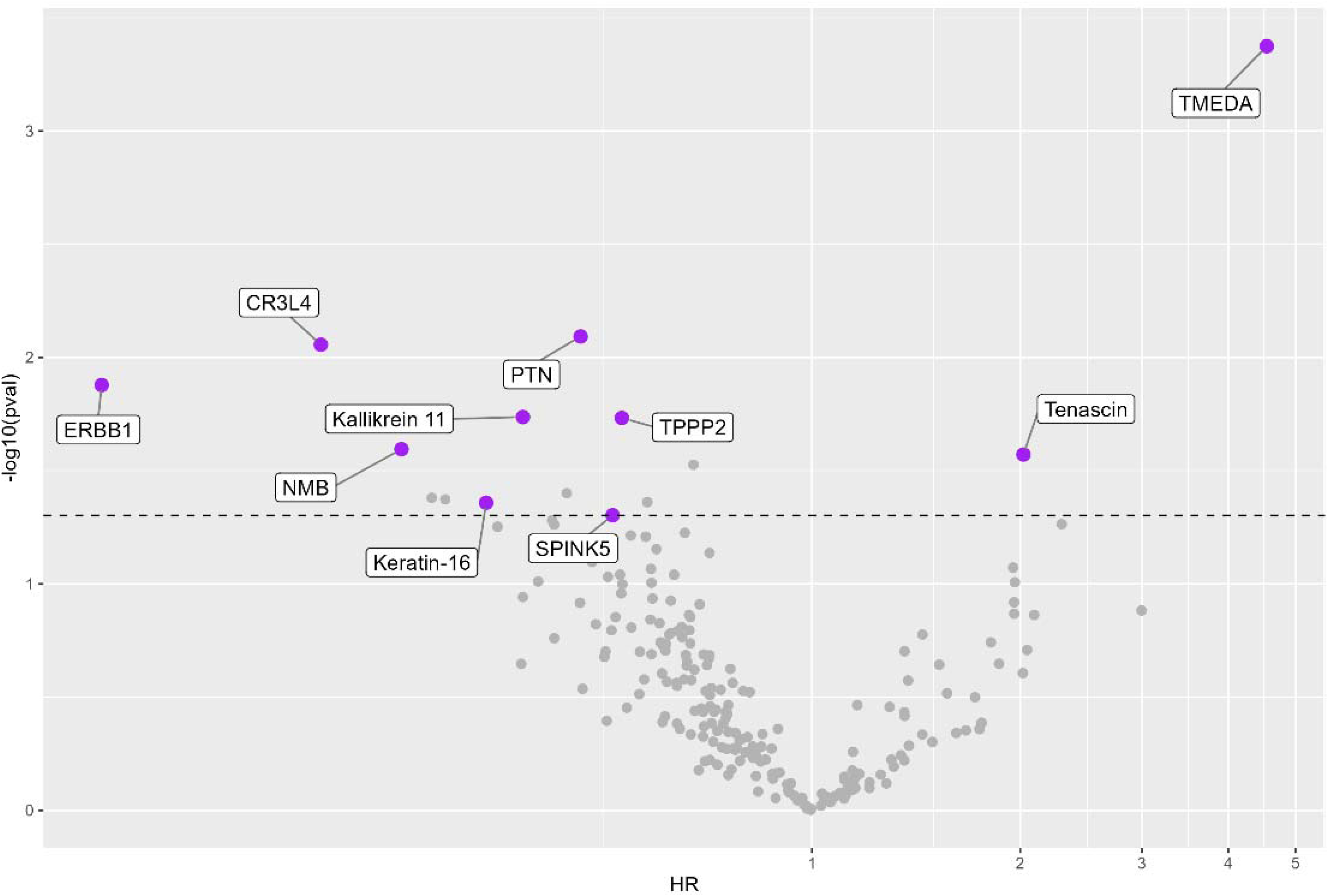
Volcano plot of association between change in protein expression and risk of heart failure hospitalization.

### Change in protein levels with EQW therapy

To understand whether the proteins we identified that differentiate EF groups are modified by EQW therapy, we then conducted linear mixed models and evaluation of interaction between treatment and timepoint on protein levels. Levels of 97 (97/221, 44%) of proteins changed from baseline to 12 months differentially by EQW treatment as compared with placebo (nominal interaction p<0.05, **Figure 3**). The minority (30/97, 31%) of these proteins were reduced to a greater degree in the EQW arm as compared with placebo; 26 of these proteins had also demonstrated baseline levels being associated with higher risk of incident HF hospitalization risk (all nominal multivariable p<0.0001) including TNC, angiopoietin-2 and NT-proBNP. TNC had also demonstrated an increased risk for hHF with increase in protein level from baseline to 12 months. These results suggest that these 26 proteins reporting at baseline on higher risk of incident hHF are beneficially modified by EQW therapy.

**Figure 3:**
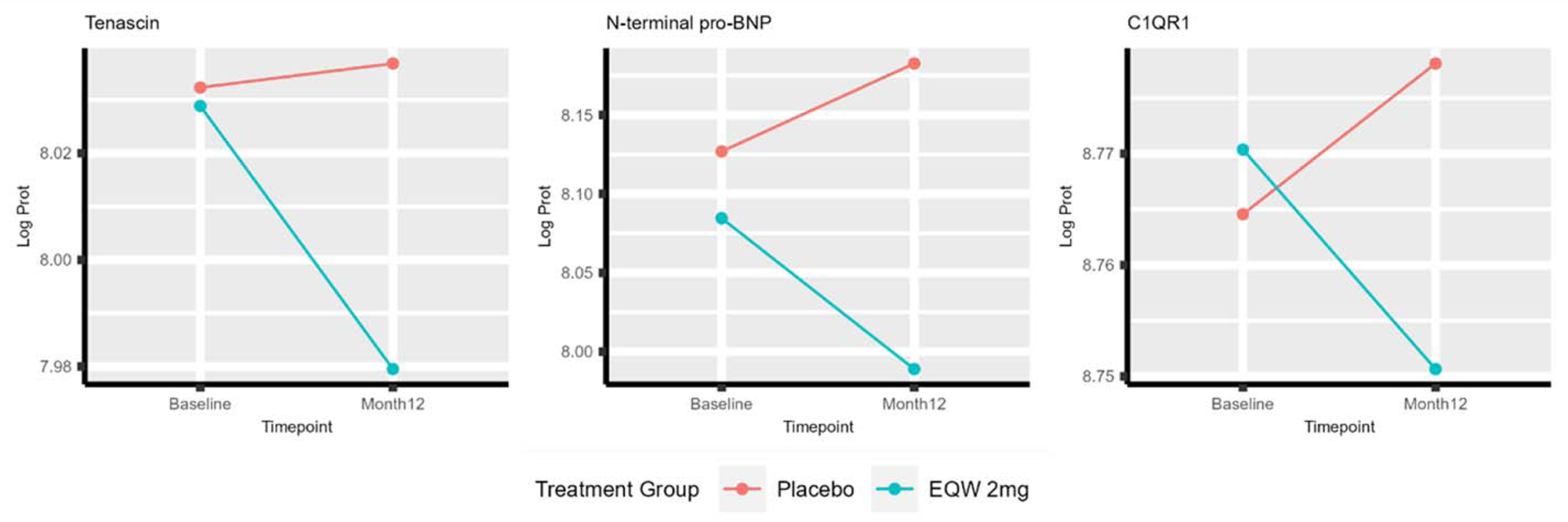
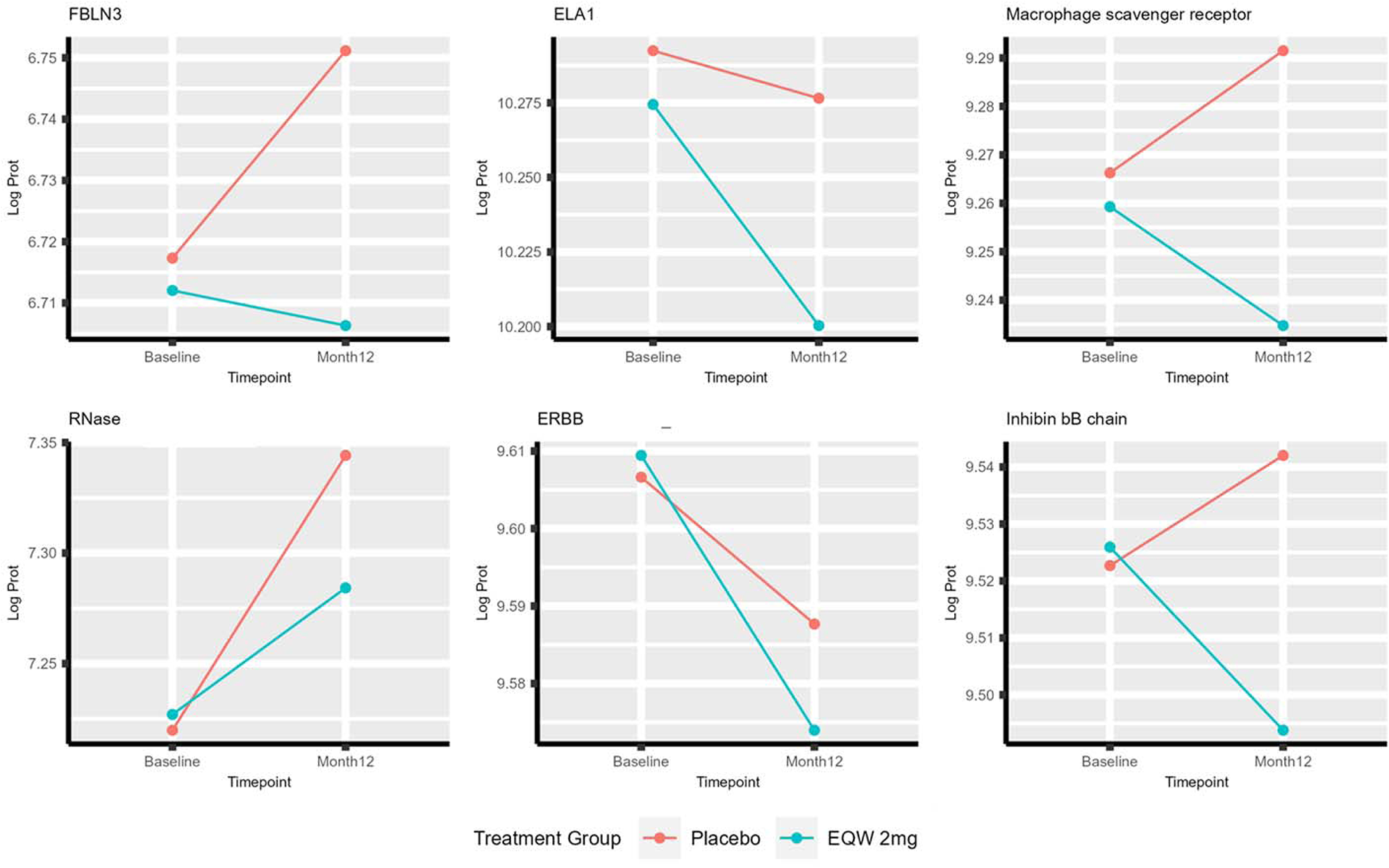
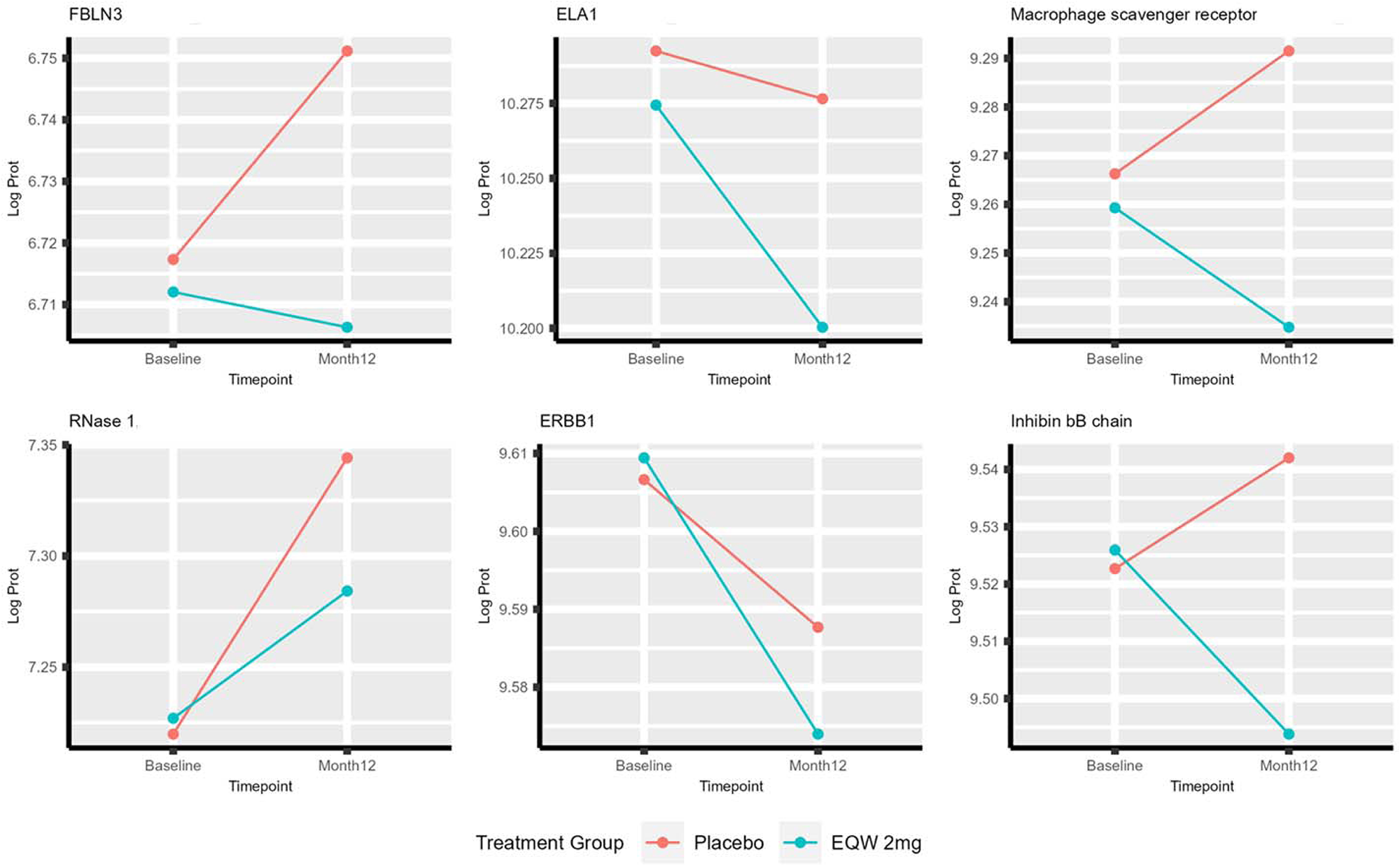
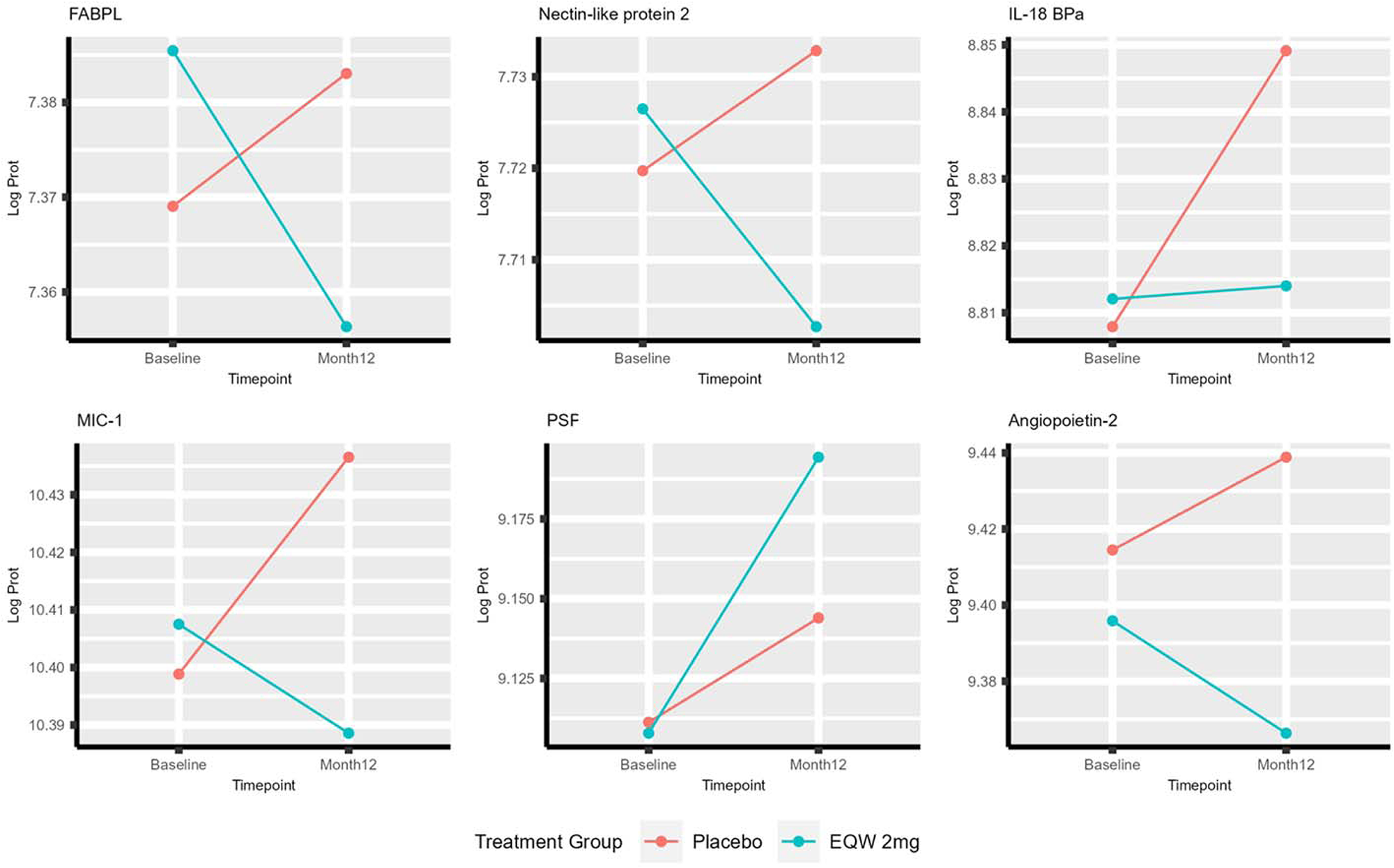
Select plots of interaction between treatment vs placebo and change in protein (baseline to 12 month) *Multiple other Somamer reagents targeting different epitopes of the Tenascin-C protein demonstrate similar patterns as displayed in the above Tenascin-C plot

**Figure 4:**
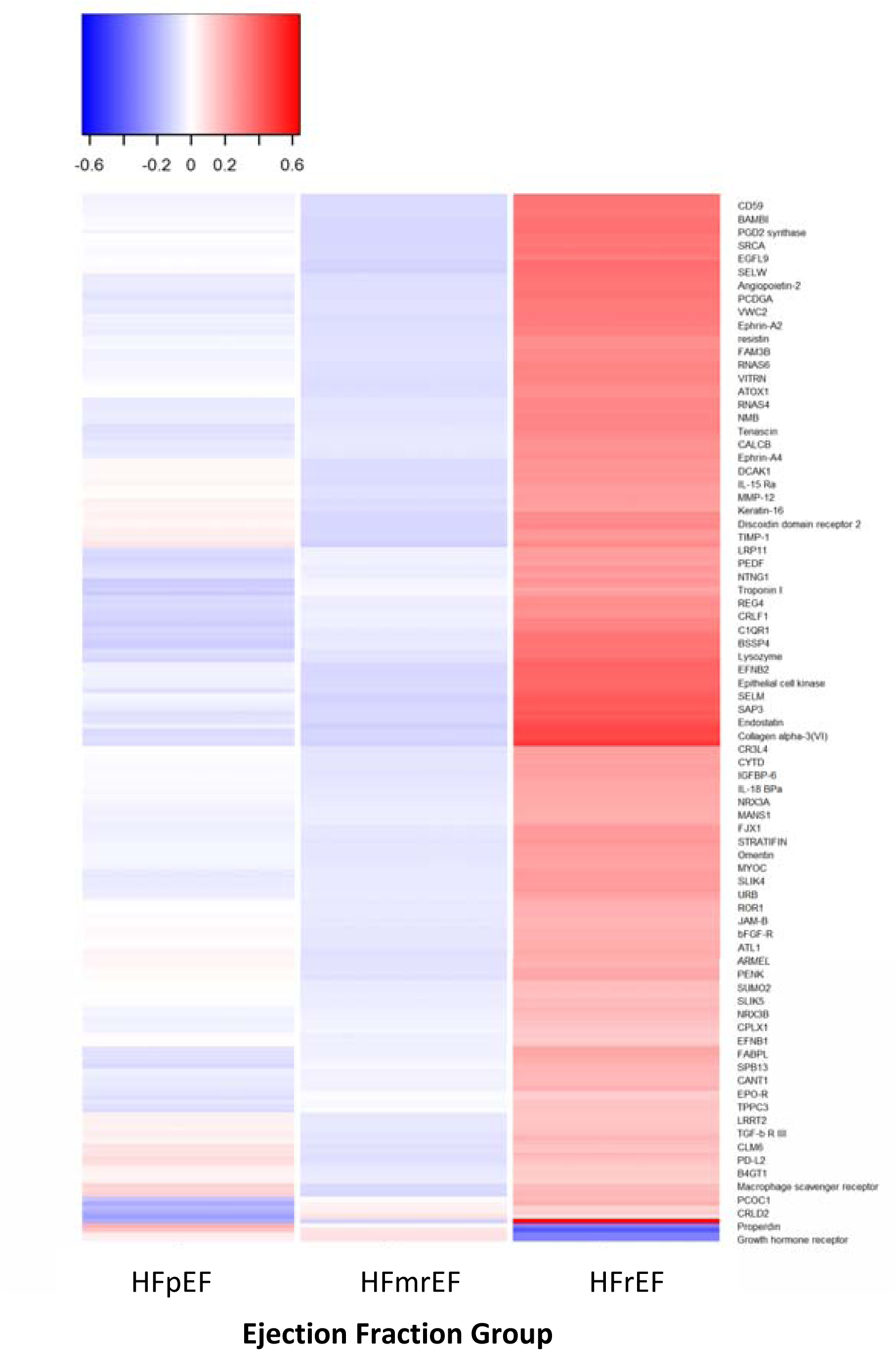
Heatmap of mean protein levels significantly different across HF groups.

## DISCUSSION

In this multinational, carefully adjudicated clinical trial cohort, we report herein the largest and most comprehensive proteomics analysis of HF across EF. Specifically, we find that the majority of proteins are concordant between HFmrEF and HFpEF. Biologic pathways represented by these proteins highlight the role of extracellular matrix regulation, angiogenesis, inflammation, and epithelial-mesenchymal transition. These findings suggest that HFmrEF may be more biologically similar to HFpEF and suggest that clinical and outcome differences seen in prior studies of HFmrEF are unrelated to underlying biologic differences.

This dominant pattern across EF groups was driven primarily by proteins with higher mean levels in individuals with HFrEF including established biomarkers such as troponin T, troponin I, angiopoietin-2, and cystatin-C (NT-proBNP was also higher in HFrEF but with a graded increase across EF as discussed below). The majority of these proteins were also prognostic for incident hHF, and several were pharmacologically modifiable by EQW (including troponin T, NT-proBNP and angiopoietin-2). Notably, the pathophysiologic domain of epithelial-mesenchymal transition (EMT), the top pathway in the overrepresentation analysis, was overexpressed in this pattern in alignment with prior HF studies in animal and translational models^32–36^ and smaller human proteomics studies.^37^ This underscores its importance in HF for the first time to our knowledge in a large, clinical cohort of human patients with HF. Tenascin C, a key extracellular matrix protein that plays a role in EMT among other processes, demonstrated this expression pattern in HFrEF compared to HFmrEF/HFpEF. Tenascin C has known pleiotropic effects but appears to primarily act as a proinflammatory and profibrotic modulator in HF, thereby worsening adverse myocardial remodeling,^38–40^ and has been shown to be elevated in both HFrEF and HFpEF.^41^ In this study, both baseline level and change in tenascin C predicted HF hospitalization and changed beneficially with EQW, demonstrating a robust profile of prognostic and modifiable biomarker that varies by EF.

Epidermal growth factor receptor (EGFR) was another protein in this dominant pattern (higher mean levels in individuals with HFrEF) that was notable throughout our analysis. Lower baseline levels of EGFR were associated with risk for hHF and decrease in EGFR from baseline to 12 months was also associated with incident hHF risk. EGFR was modified by EQW therapy, but in a potentially unfavorable direction (decreased in intervention arm). These data support further investigation into the role of EGFs and EGFRs in heart failure, a pathophysiologic phenomenon that appears complex and incompletely understood to date. While a degree of EGF/EGFR activity is normal for cardiac development and functioning,^42, 43^ overexpression of EGF may be associated with CV disease, atrial fibrillation, and HF.^42–44^ Further, EGFR overexpression in mouse models may promote HOCM-like phenotypes with regression of this phenotype by EGFR inhibition.^45^ On the other hand, blocking EGFR signaling with tyrosine kinase inhibition in humans is associated with development of HF in rare cases.^46^ Findings from the present study are unable to determine causality or fully elucidate the role of EGFR signaling, but support the prognostic role of EGFR in patients with HF, particularly those with HFmrEF/HFpEF.

The second most common pattern was a graded progression across EF groups with significant differences between HFrEF, HFmrEF, and HFpEF. This included the established biomarker, NTpro-BNP, as well as proteins of immune regulation (properdin, hepatitis A virus cellular receptor 2) and those reflecting renal injury (e.g. neutrophil gelatinase-associated lipocalin, a previously recognized biomarker with mixed evidence in HF^47–50^) among others. Properdin was the only protein to demonstrate the relationship of lower levels in HFrEF, which were associated with increased risk for adverse outcomes (consistent with prior studies)^51^, and a graded increase in level as EF increased. The other 5 proteins were higher in HFrEF and each of these proteins were also associated with hHF risk, except for transcobalamin-1.

Lastly, several proteins (n=3) involved in redox homeostasis/antioxidant processes (selenoprotein W) and immune regulation (CD48 antigen and programmed cell death 1 ligand 2) were significantly *underexpressed* in HFmrEF compared to HFrEF/HFpEF, and lower levels were associated with lower risk for subsequent hHF. Additionally, three proteins demonstrated significantly higher levels in both HFmrEF and HFrEF compared to HFpEF including prokineticin-2, which was associated with increased hHF risk at higher levels in this study. In addition to its neurologic and gastrointestinal effects, prokineticin-2 has also been shown to play a role in cardiac hypertrophy in hypertensive pressure-overload mice models.^52^ Although prokineticin-2 was not differentially reduced by EQW treatment in this study, this protein pathway may warrant further investigation in HF pathophysiology.

This study also adds to the growing literature describing biologic mechanisms leading to the development and progression of HF. These previous studies have identified dominant biologic domains with considerable overlap to the present study including the domains of inflammation/apoptosis and extracellular matrix remodeling (and related angiogenesis processes).^53–56^ Further, this analysis extends and builds upon findings from similar prior studies studying the biology of the EF spectrum.^4, 5, 13, 14, 41, 57^ A few early studies suggested that HFmrEF may have an intermediate biomarker profile between HFrEF and HFpEF.^5, 14^ For instance, one study demonstrated the top upregulated pathways in HFmrEF were related to neutrophil degranulation, leucocyte migration, and DNA-binding transcription factor activity, and this was intermediate between the dominant biological pathways of HFrEF and HFpEF.^14^ Another study found only BNP, KIM-1, RBC, and Hgb significantly associated with EF and intermediate in HFmrEF.^5^ These analyses had several limitations including being based on a relatively small number of proteins (<100).^5, 14^

Consistent with our findings, a more recent study analyzing 1129 proteins using the SomaLogic platform in 173 patients found that HFmrEF was more biologically similar to HFpEF, although the degree of this association was affected by whether patients have HFmrEF with recovered EF (i.e. previously HFrEF) or unchanged/stable EF.^4^ The analysis noted several differences in biological themes across LVEF groups such as within the domain of ‘VEGF A/angiogenesis’ including the proteins angiopoietin-2 and VEGF-A and within the domain of growth factor signaling including insulin-like growth factor-binding proteins. Each of these signals was overexpressed in HFrEF and similar in HFmrEF and HFpEF,^4^ which is consistent with our findings. On the other hand, the analysis also noted prominent MMP-activity differentially in HFrEF patients,^4^ which was not as notable in our study. This analysis also described a few overexpression signals in HFmrEF (complement/opsonization related) and separately in HFpEF (NK-cell markers, VEGF-C/angiogenesis),^4^ which were not noted in our study; still, these signals were fewer than in HFrEF, consistent with our results. Taken together, the findings of our present study and prior analyses suggest that HFmrEF may be a heterogenous biological entity (partly driven by preceding EF trend) but that patients with HFmrEF predominantly demonstrate biological similarities to those with HFpEF.

The results of the present analysis also suggest that unique biologic patterns associated with HFmrEF, HFpEF, or HFmrEF/HFpEF (e.g. EF>40%) compared to HFrEF are fairly narrow (<10 uniquely elevated proteins). It should be emphasized that this result does not indicate that these biologic processes (e.g. ECM regulation, endothelial function/angiogenesis, inflammation) are not present in HFmrEF/HFpEF compared to non-HF patients; several prior analyses have demonstrated the role of these processes in HFmrEF/HFpEF.^58–62^ But rather, the present analysis finding indicates these processes (as reflected by systemic protein expression) are more highly expressed in HFrEF and concomitant DM as compared to HFmrEF/HFpEF and concomitant DM. Several factors may explain this finding. First, HFmrEF/HFpEF is certainly a heterogenous entity and may be sufficiently heterogenous, when assessed by clinical HF status (as opposed to hemodynamically, for instance), to diminish unique biologic signals. This analysis’ cohort likely includes HF with recovered EF, low risk early HFpEF, and HF with declining EF (55% to 45%) along with a range of HFpEF phenotypes. Second, the HF cohort in this diabetes trial may be artificially distinct from routinely encountered patients with HF; for instance, HFpEF patients are typically older with higher burden of DM compared to patients with HFrEF^12^ and, in this study, HF patients across EF spectrum were similarly aged and all had DM as per inclusion criteria. Third, the strength of the signal for protein elevations in the HFrEF group may have been skewed by patients with severely reduced EF (n=45/211). Fourth, residual confounding by factors unadjusted for in the analyses, such as renal function or burden of ischemic vs non ischemic cardiomyopathy, may have affected the strength of protein elevations in the HFrEF cohort. Still, it should be noted that the degree of rigor in covariable adjustment is higher in the present study than in previous gold standard studies in this area.^4, 14^ Within the context of these possible limitations, this study’s findings suggest that EF, when applied to a broad chronic HF population, has limited biologic specificity as EF increases above 40% (only ∼2% of proteins with unique overexpression in HFmrEF/HFpEF). Taken together, these findings support this movement to expand HF phenotyping and classification beyond EF, particularly when considering patients with EF >40%, to inform development and testing of targeted therapies.

### Strengths and Limitations

Notably, the present study included the largest combination of cohort sample size (1,199 participants) and proteomics panel scale (4,979 proteins). Further, to our knowledge, this is the first study to combine this scale of biologic investigation and include prognostication by *change in protein* levels across three EF groups with additional interaction analyses by treatment (EQW). However, it is important to note several limitations of our study. First, we used EF categorization as collected and documented in the parent EXSCEL trial, which defined a ‘mid range’ EF as 40-55%, since individual level EF results were not available; these EF strata differ slightly from the HF EF categories more recently established since the initiation of the EXSCEL trial, including the category of HFmrEF with EF 41-49%.^1, 2^ However, the variability of EF assessment makes it unlikely that such slight changes in EF thresholds would dramatically alter protein patterns evaluated on a large-scale as in this study. Serial values of ejection fraction were not available in EXSCEL so we were unable to assess sub-categories of HFmrEF/HFpEF recovered vs. unchanged. EXSCEL enrolled participants with DM with a high proportion with prevalent CAD, thus our results may not be generalization to other populations with HF. Finally, we used a broad HF definition; protein differences might be more manifest in a more strictly defined HFmrEF/HFpEF population (i.e. with additional requirements of natriuretic peptide elevation and/or invasively confirmed elevated filling pressures).

## CONCLUSIONS

In this large clinical trial, we found that most proteins were similar between HFmrEF and HFpEF and differentially elevated in HFrEF in patients with T2DM. These results suggest that HFmrEF is more biologically similar to HFpEF and highlight prognostic and modifiable biomarkers that vary by EF.

## Data Availability

Due to its proprietary nature and confidentiality agreements, supporting data cannot be made openly available.

## ACKNOWLEDGEMENTS

We thank the participants of the EXSCEL clinical trial for their participation.

---

**Central Illustration:**
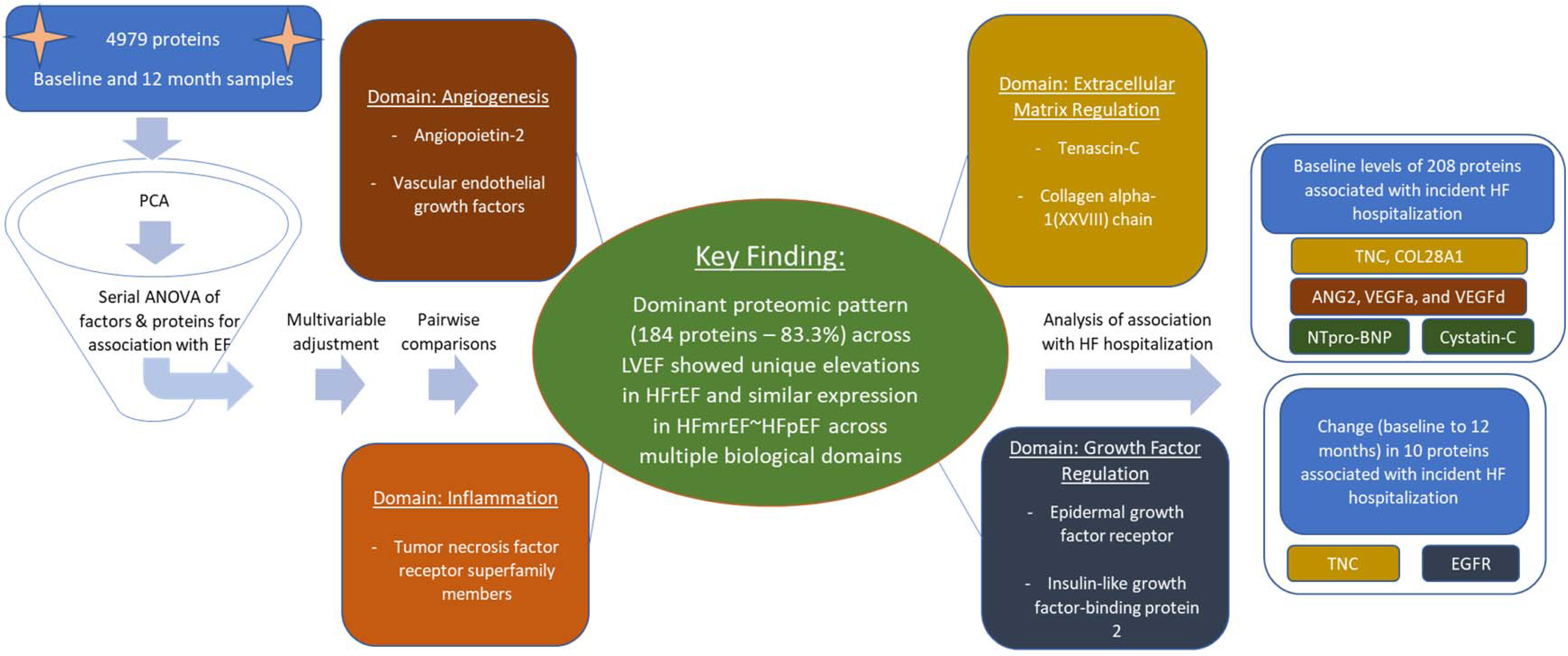
Study flowchart and key findings.

